# Plasma metabolomic profiling of frailty in older adults at risk of dementia

**DOI:** 10.64898/2026.10.06.26364906

**Authors:** Isabelle Alldritt, Johannes C. Michaelian, Rachael Yu, Sarah E. Deemer, Richard J. Mills, Sharon L. Naismith, Andrew Philp

**Author notes:** Joint first authors (Isabelle Alldritt, Johannes C. Michaelian). Joint senior authors (Sharon L. Naismith, Andrew Philp).

## Abstract

**Background:** Frailty is a prominent risk factor for dementia. Early detection of at-risk individuals may enable intervention to prevent or slow dementia pathology. Blood-based biomarker discovery could offer a path for early detection of frailty and cognitive decline.

**Methods:** Older adults at risk of dementia underwent comprehensive multi-disciplinary assessment at the Healthy Brain Ageing (HBA) clinic, including geriatric, neurophysiological and mood evaluations, in addition to providing a fasted blood sample. Clinical data were used to calculate a deficit accumulation frailty index (FI) score. Additional cohorts attended either one (n=118) or two (n=73) subsequent visits over an average period of 4 years. Untargeted metabolomics was performed on plasma samples, and metabolites were clustered by similarity. Linear regression was used to assess association of clusters with FI. Finally, change in plasma abundance over time was assessed.

**Results:** At assessment, participants had a mean (SD) age of 68.10 (8.07) years; 72.2% were female. Hierarchical clustering identified 10 clusters of metabolites, of which 6 were significantly associated with FI (p<0.05) at baseline and contained similar classes of metabolite. Differential abundance analysis found unique temporal regulation of metabolite abundance in each cluster, but across clusters severity of frailty impacted direction of change in plasma abundance over time.

**Conclusions:** Frailty index was associated with carboxylic acids and derivatives, fatty acyls and organooxygen compounds across multiple clusters, and a large number of lipids were linked to frailty score, potentially implicating dysregulation of energy metabolism in the pathophysiology of frailty and forming a panel of predictive biomarkers of at risk patients.

## Introduction

The World Health Organisation predicts that the population of people aged 60 or older will reach 1.4 billion in 2030 (1). With an ageing global population, the impact of comorbidities of ageing are becoming increasingly relevant. Age is the single greatest risk factor for cognitive decline, which can range from subjective cognitive decline (SCD) or mild cognitive impairment (MCI) to dementia. Dementia affected an estimated 433,300 Australians in 2025 and is the second leading cause of death in Australia (2). Globally, it is estimated that there are 10 million new cases of dementia per year, representing the seventh leading cause of death (3). However, despite high prevalence, it can take years to receive a diagnosis of dementia after initial presentation of symptoms (4,5) with an estimated 50% of adults with cognitive impairment receiving inappropriate clinical evaluation (6).

Older adults living with SCD and MCI are at risk of dementia, however these clinical states do not necessarily translate to a full dementia prognosis. For example, the annual rate of progression of MCI to dementia ranges from approximately 5 to 17% (7). One study found that over a 4 year period, only 7% of individuals with SCD progressed to dementia (8), while a recent systematic review calculated the annual conversion rate for dementia in individuals with SCD to be 1.12% (9). The pathology of dementia can begin decades before the clinical manifestation of symptoms, with recent evidence suggesting early intervention can prevent or delay approximately 45% of dementia occurance (10).

One clinical factor that may classify individuals as high risk of dementia is physical frailty. Frailty is a complex, multifactorial condition which causes a state of increased vulnerability, underlined by a decline in reserve and function across multiple physiological systems. Although there is still no consensus clinical definition of frailty, two approaches are commonly used to classify frail individuals: the frailty phenotype (FP) and frailty index (FI). The FP characterises frailty through an set of observable patient characteristics (11), whereas the FI quantifies level of frailty using a ‘cumulative deficit’ model (12). Although robust association between dementia, FP or FI have been reported, the association between frailty and SCD or MCI is less well characterised.

There is an increasing interest in identifying biomarkers of frailty in individuals with or at risk of dementia, although none have yet been conclusively established. For instance, a recent cross-sectional analysis found frail individuals demonstrated lower global cognition and had significantly higher plasma levels of neurofilament light chain (NfL), suggesting an association with cognitive ability and general neurodegenerative processes (13). A systematic review of the current literature found five biomarkers, including NfL, were significantly associated with both frailty and cognition, however the authors identified that many studies may exclude MCI due to lack of neuropsychiatric testing (14). Additionally, blood-based biomarkers associated with amyloid and tau pathologies (phosphorylated tau, amyloid beta ratios or NfL) have shown correlations with brain deposits of amyloid and tau and accuracy in predicting incident dementia in clinical settings (15). However, their capacity as screening tools in community settings is not entirely clear (16), and the exclusive use of a limited set of late stage markers may exclude detection of broader changes in metabolic signalling networks occurring at early stages of or preceding dementia pathophysiology.

Untargeted metabolomics is a highly sensitive, unbiased profiling technique that can provide a comprehensive overview of molecular signatures relevant to a given disease or condition There has been growing interest in metabolomics as a technique for studying dementia. For instance, dysregulation in serum amino acids in individuals with subjective memory complaints and MCI which distinguishes them from healthy controls has previously been observed (17). Untargeted metabolomics has also identified decreased benzoic and hippuric acid levels in MCI participants compared to those with Alzheimer’s disease, and a pattern of disturbed lipid and amino acid metabolism in all participants with cognitive decline compared to healthy controls (18). However, neither study considered the presence of clinical comorbidities, such as frailty, which increase the risk of dementia in MCI populations. As such, unbiased investigations into frail clinical populations with SCD or MCI who are the most at risk of developing dementia are lacking.

Therefore, in this study, we utilised mass spectrometry based untargeted metabolomics to profile the plasma metabolome of 212 patients at risk of dementia. Our goal was to identify circulating metabolomic markers of frailty that may provide a link between increased frailty and dementia risk.

## Methods

### Participant recruitment

Participants were recruited from the Healthy Brain Ageing (HBA) research clinic at the Brain and Mind Centre, The University of Sydney between 2020 and 2023. Ethical approval for the project Healthy Brain Ageing Program: ‘Discovering Novel Biomarkers and Interventions’ was obtained from the University of Sydney Human Research Ethics Committee (identifier 2019/HE000271). All participants provided written informed consent prior to participating in the study. All data was obtained in compliance with the Declaration of Helsinki. A subset of participants subsequently attended follow up visits to the research clinic on average 2.16 years (±1 year) and 3.91 years (±0.7 years) after baseline visit.

Demographic and clinical variables of interest included age, sex, educational level, body mass index (BMI), living circumstances, activities of daily living, and measures of anxiety and depression. In addition, Apolipoprotein E4 (APO 4) carrier status and estimated glomerular filtration rate (eGFR) were determined and used as covariates.

### Calculation of frailty index

Frailty was operationalised using a deficient accumulation approach, consistent with the methodology described by Searle and colleagues (19). A total of 30 health-related variables, selected according to established criteria (19), were included in the calculation of FI score and applied uniformly across participants.

Variables were incorporated from four broad domains: medical history and comorbidities, anthropomorphic measures, organ system disease burden assessed using the Cumulative Illness Rating Scale-Geriatric, and mood and self-reported health. Full details of collected variables are reported in eTable 1 in the Supplement. Together, these variables provided broad coverage of chronic disease burden, metabolic and cardiovascular health, neurological and psychiatric morbidity, and global health perception.

Each deficit was coded on a continuous scale from 0 to 1, where higher values indicated greater deficit severity. Binary variables were coded as 0 (absence of the deficit) or 1 (presence of the deficit). Ordinal variables were graded according to severity, with intermediate values reflecting partial expression of a deficit (e.g. 0.5).

Participants were excluded from frailty index analyses if they had more than five missing deficits (> 5/30 variables). For participants with five or fewer missing values, missing deficits were imputed using the sample mean for that variable, consistent with common practice in frailty index construction. All deficits were weighted equally. The frailty index for each participant was calculated as the sum of deficit scores divided by the total number of deficits considered: Frailty Index = Sum of deficits present/number of deficits included. Resulting frailty index scores ranged from 0 to 1, with higher values indicating greater accumulation of health deficits and increased frailty. The full list of deficits included are provided in eTable 1 in the Supplement. As FI cutoffs to define frailty are study population-specific, we grouped participants into 4 quartiles of increasing levels of frailty: quartile 1 (0<FI<0.1379), quartile 2 (0.1400<FI<0.1810), quartile 3 (0.1811<FI<0.2586), and quartile 4 (0.2587<FI<0.5086).

### Blood collection and processing

Following an overnight fast, venous blood samples were obtained on the morning of the clinical assessment using ethylenediaminetetraacetic acid (EDTA) vacuum and serum separating tubes (SST) tubes. Plasma collected in EDTA tubes was centrifuged within 30 minutes of collection, while SST-serum samples were left at room temperature for approximately 30 minutes to allow clot formation before centrifuged. Both EDTA and SST tubes were centrifuged at 1,800 × g for 10 minutes at 4 °C, after which the resulting plasma and serum aliquots were immediately stored at –80 °C until analysis.

### APOE genotyping

Genomic DNA used for APOE genotyping was extracted from peripheral blood mononuclear cells and analysed using the UK Biobank Axiom™ Array. Genotyping was performed at the Ramaciotti Centre for Genomics, UNSW Australia (NATA ISO/IEC Accreditation #20417), using the same array to identify the APOE single nucleotide polymorphisms (SNPs) rs429358 and rs7412. Based on these variants, participants were categorised according to APOE-ε4 status, with carriers defined as individuals possessing at least one ε4 allele and non-carriers defined as those without an ε4 allele.

### Metabolomics

Metabolites were extracted from 50µl plasma using a 1:6.4:6.4:4.76 ratio of methanol, chloroform and water (20). Samples were agitated at room temperature for 2 minutes, incubated at 4°C for 10 minutes, and centrifuged at 17,000 xg for 10 minutes at 4°C. Polar and non-polar phases were removed to separate Eppendorf tubes and dried under a vacuum. Prior to analysis, samples were suspended in 75:25 MeCN:ddH2O and 75:25 IPA:ddH2O for HILIC and reverse phase chromatography, respectively. 10 µl was removed from each suspended plasma sample to a clean Eppendorf tube to create a representative quality control (QC) sample.

Metabolomics analysis was performed on an Ultimate 3000 UHPLC pump and autosampler coupled to a Q-Exactive mass spectrometer (Thermo Fisher Scientific, USA) operated in positive and negative ionisation modes. For HILIC chromatography, an InfinityLab Poroshell 120 HILIC-Z column (Agilent) was used with mobile phase A 10mM ammonium formate in 95% acetonitrile with 0.1% formic acid and mobile phase B 10mM ammonium formate in 50% acetonitrile with 0.1% formic acid. For reverse phase chromatography, a Zorbax SB-Aq RRHD column (Agilent) was used with mobile phase A ddH2O with 0.1% formic acid and mobile phase B methanol with 0.1% formic acid. 5µl sample was injected. Injection order was randomised to avoid bias. 8 pooled QC samples were injected at the start of the run for column conditioning, followed by an injection every 10 biological samples.

Data was converted to an open mzXML format using MS Convert (ProteoWizard, USA) and processed using inhouse R scripts (21) utilising the XCMS package (22). Chromatograms were extracted using the centWave algorithm. Tolerance was set at 20ppm. Retention time alignment was performed using the Obiwarp method with a bin size of 0.6. Bandwidth parameters for correspondence analysis were determined manually for each ion mode and polarity. Following correspondence, missing values were imputed via integration of peak areas.

Following recommendation guidelines for untargeted metabolomics (23) metabolite features were retained when peaks were present in at least 70% of QC samples, relative standard deviation was less than 30%, and the extraction blank to mean QC peak area was less than 50%. Sample drift was corrected using probabilistic quotient normalisation, then missing values were imputed using a Random Forest algorithm. Data were log transformed prior to analysis.

### Statistical analysis

Statistical analysis was performed in R v4.5.2. Using baseline samples to reduce intra-individual variation, metabolites were assigned to clusters by hierarchical clustering. Principal component analysis (PCA) was performed to calculate a cluster membership score. To determine association between metabolite clusters and frailty index, three regression analyses were performed. The first model used an unadjusted linear model, with FI set as the independent variable and cluster score set as the dependent variable. Next, the model was adjusted for age in years and sex. Finally, the model was adjusted for age in years, age in years^2^, sex, eGFR, APOE ε4 allele carrier status, years of education, cognitive diagnosis and MMSE score. The analysis was performed separately on polar and non-polar metabolites acquired from positive and negative ionisation modes.

Metabolites were identified using the metid package (24) against publicly available datasets downloaded from HMDB and KEGG websites. Over representation analysis was performed using MetaboAnalyst 5.0 (25) to assign identify enriched metabolite classes per cluster.

To examine change in metabolite abundance between baseline and subsequent visits, p-values were calculated by a linear model constructed using limma (26). Comparisons were made between baseline and the initial follow up visit in the lowest quartile and highest quartile individually, then an interaction comparison was fitted to assess which metabolites respond differently over time in the highest quartile relative to the lowest quartile. To account for false discovery rate, all p-values were adjusted using the Benjamini-Hochberg method. Significance was set at p*_adj_*<0.05.

## Results

### Participant characteristics

At the baseline visit (n=212), mean age was 68.10 years (±8.07), with 72.2% of participants female. 76 participants were classified as SCD and 136 were classified as MCI. Participants were divided into quartiles by their calculated FI score (quartile 1, n=55; quartile 2, n=53; quartile 3, n=52; quartile 4, n=52), with quartile 1 (Q1) having the lowest FI score and quartile 4 (Q4) having the highest FI score.

At the first follow up visit (LF1, n = 118), mean age was 68.39 years (±7.94) and 70.3% were female. At the second follow up visit (LF2, n=73), mean age was 71.52 years (±7.20) and 65.8% were female. Detailed participant characteristics are provided in Table 1. Number and characterisation of quartiles at baseline are provided in Table 2.

**Table 1.** Participant characteristics across HBA cohort

|  | **Baseline (BL)** | **2-year follow up (LF1)** | **4-year follow up (LF2)** |
| --- | --- | --- | --- |
| **Number of participants (n)** | 212 | 118 | 73 |
| **Age (years)** | 68.10±8.07 | 68.39±7.94 | 71.52±7.20 |
| **Sex, n (%)** |  |  |  |
| Male | 59 (39) | 35 (30) | 25 (34) |
| Female | 153 (61) | 83 (70) | 48 (66) |
| **Education (years)** | 14.38±2.97 | 14.29±2.74 | 14.42±2.90 |
| **ADL score** | 16.97±0.29 | NA | NA |
| **Cognitive diagnosis** |  |  |  |
| MCI (yes/no) | 136/76 | 46/75 | 29/46 |
| aMCI, n (%) | 69 (51) | 19 (41) | 10 (34) |
| naMCI, n (%) | 67 (49) | 27 (59) | 19 (66) |
| **MMSE** | 28.41±2.06 | 28.79±2.05 | 28.90±1.46 |
| **CIRS-G** |  |  |  |
| Total score | 5.95±3.26 | 4.63±3.17 | 4.07±3.09 |
| Severity index | 1.35±0.31 | 1.40±0.44 | 1.45±0.48 |
| **Geriatric Anxiety Inventory** | 4.87±5.32 | 4.92±5.54 | 3.52±3.67 |
| **Geriatric Depression Scale** | 3.84±3.74 | 3.58±3.57 | 2.76±2.35 |
| **ApoE4 carrier (n)** | 40 | 24 | 19 |
| **ApoE genotype (n)** |  |  |  |
| Homozygote | 2 | 2 | 0 |
| Heterozygote | 38 | 22 | 19 |
| Non-carrier | 81 | 52 | 32 |
| Not available | 91 | 45 | 24 |

Note. Unless otherwise specified, data are presented as mean ± standard deviation.

**Table 2.**
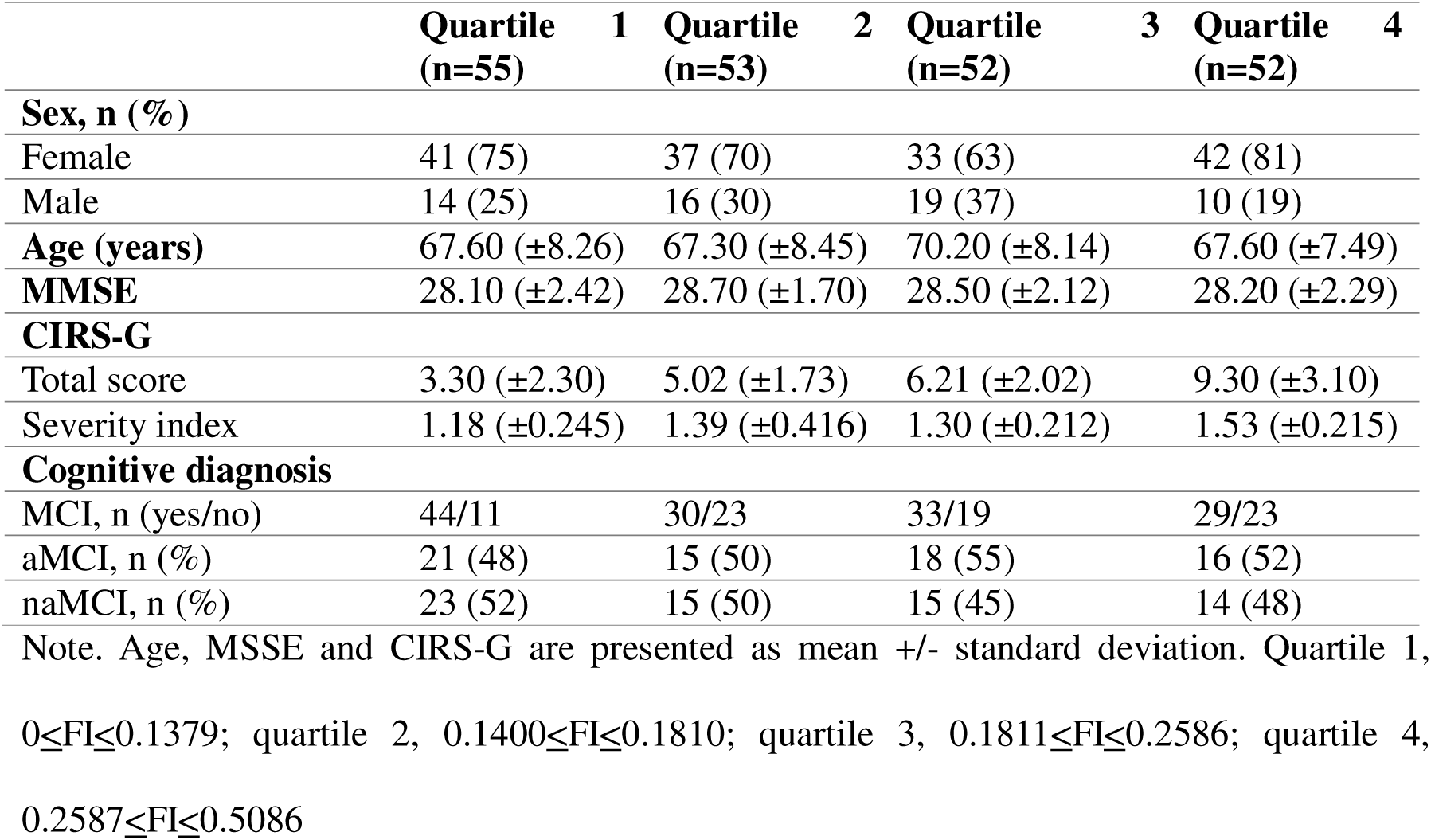
Characteristics of participants at baseline based on frailty index quartiles

|  | **Quartile 1**<br>**(n=55)** | **Quartile 2**<br>**(n=53)** | **Quartile 3**<br>**(n=52)** | **Quartile 4**<br>**(n=52)** |
| --- | --- | --- | --- | --- |
| **Sex, n (%)** |  |  |  |  |
| Female | 41 (75) | 37 (70) | 33 (63) | 42 (81) |
| Male | 14 (25) | 16 (30) | 19 (37) | 10 (19) |
| **Age (years)** | 67.60 (±8.26) | 67.30 (±8.45) | 70.20 (±8.14) | 67.60 (±7.49) |
| **MMSE** | 28.10 (±2.42) | 28.70 (±1.70) | 28.50 (±2.12) | 28.20 (±2.29) |
| **CIRS-G** |  |  |  |  |
| Total score | 3.30 (±2.30) | 5.02 (±1.73) | 6.21 (±2.02) | 9.30 (±3.10) |
| Severity index | 1.18 (±0.245) | 1.39 (±0.416) | 1.30 (±0.212) | 1.53 (±0.215) |
| **Cognitive diagnosis** |  |  |  |  |
| MCI, n (yes/no) | 44/11 | 30/23 | 33/19 | 29/23 |
| aMCI, n (%) | 21 (48) | 15 (50) | 18 (55) | 16 (52) |
| naMCI, n (%) | 23 (52) | 15 (50) | 15 (45) | 14 (48) |

Note. Age, MSSE and CIRS-G are presented as mean +/- standard deviation. Quartile 1, $0 \leq FI \leq 0.1379$ ; quartile 2, $0.1400 \leq FI \leq 0.1810$ ; quartile 3, $0.1811 \leq FI \leq 0.2586$ ; quartile 4, $0.2587 \leq FI \leq 0.5086$

**Table 3.** Linear regression of frailty index against cluster membership score

|  | **$\beta$ [95% CI]** | **p** | **R<sup>2</sup>** |
| --- | --- | --- | --- |
| **Model 1** |  |  |  |
| **Cluster 1** | 0.006<br>[0.000, 0.012] | 0.037 | 0.024 |
| **Cluster 2** | 0.017<br>[0.004, 0.030] | 0.013 | 0.033 |
| **Cluster 3** | -0.001<br>[-0.002, -0.000] | 0.027 | 0.033 |
| **Cluster 4** | 0.002<br>[0.000, 0.003] | 0.018 | 0.037 |
| **Cluster 5** | -0.006<br>[-0.013, -0.000] | 0.037 | 0.029 |
| **Cluster 6** | -0.004<br>[-0.008, -0.001] | 0.025 | 0.033 |
| **Cluster 7** | 0.009<br>[0.001, 0.017] | 0.026 | 0.027 |
| **Cluster 8** | -0.009<br>[-0.017, -0.000] | 0.044 | 0.022 |
| **Model 2** |  |  |  |
| **Cluster 1** | 0.006<br>[0.001, 0.012] | 0.023 | 0.038 |
| **Cluster 2** | 0.022<br>[0.009, 0.034] | 0.001 | 0.068 |
| **Cluster 3** | -0.001<br>[-0.002, -0.000] | 0.007 | 0.058 |
| **Cluster 4** | 0.002<br>[0.000, 0.003] | 0.008 | 0.057 |
| **Cluster 5** | -0.007<br>[-0.013, -0.001] | 0.015 | 0.050 |
| **Cluster 6** | -0.004<br>[-0.007, -0.001] | 0.016 | 0.049 |
| **Model 3** |  |  |  |
| **Cluster 1** | 0.006<br>[0.001, 0.011] | 0.028 | 0.164 |
| **Cluster 2** | 0.017<br>[0.005, 0.030] | 0.008 | 0.175 |
| **Cluster 3** | -0.001<br>[-0.002, -0.000] | 0.018 | 0.139 |
| **Cluster 4** | 0.002<br>[0.000, 0.003] | 0.014 | 0.142 |
| **Cluster 5** | -0.007<br>[-0.013, -0.001] | 0.024 | 0.135 |
| **Cluster 6** | -0.004<br>[-0.007, -0.001] | 0.021 | 0.137 |

Note. Model 1 was unadjusted. Model 2 was adjusted for age and sex. Model 3 was adjusted for age, age<sup>2</sup>, sex, eGFR, ApoE4 allele carrier status, years of education, cognitive diagnosis and MMSE score.

### Hierarchical clustering of metabolites

Untargeted metabolomics analysis was performed on 403 plasma samples. After quality control procedures were performed, a total of 2403 polar and 855 non-polar metabolite features were retained for analysis. We first performed exploratory analysis to group metabolites by similarity. Data was stratified by timepoint, and hierarchical clustering was performed on baseline samples of each polarity and ionisation, identifying 10 clusters in both polar and non-polar datasets (Figure 1A-D). Within each cluster, a cluster membership score was generated for each metabolite using PCA which was used to assess association with FI.

**Figure 1.**
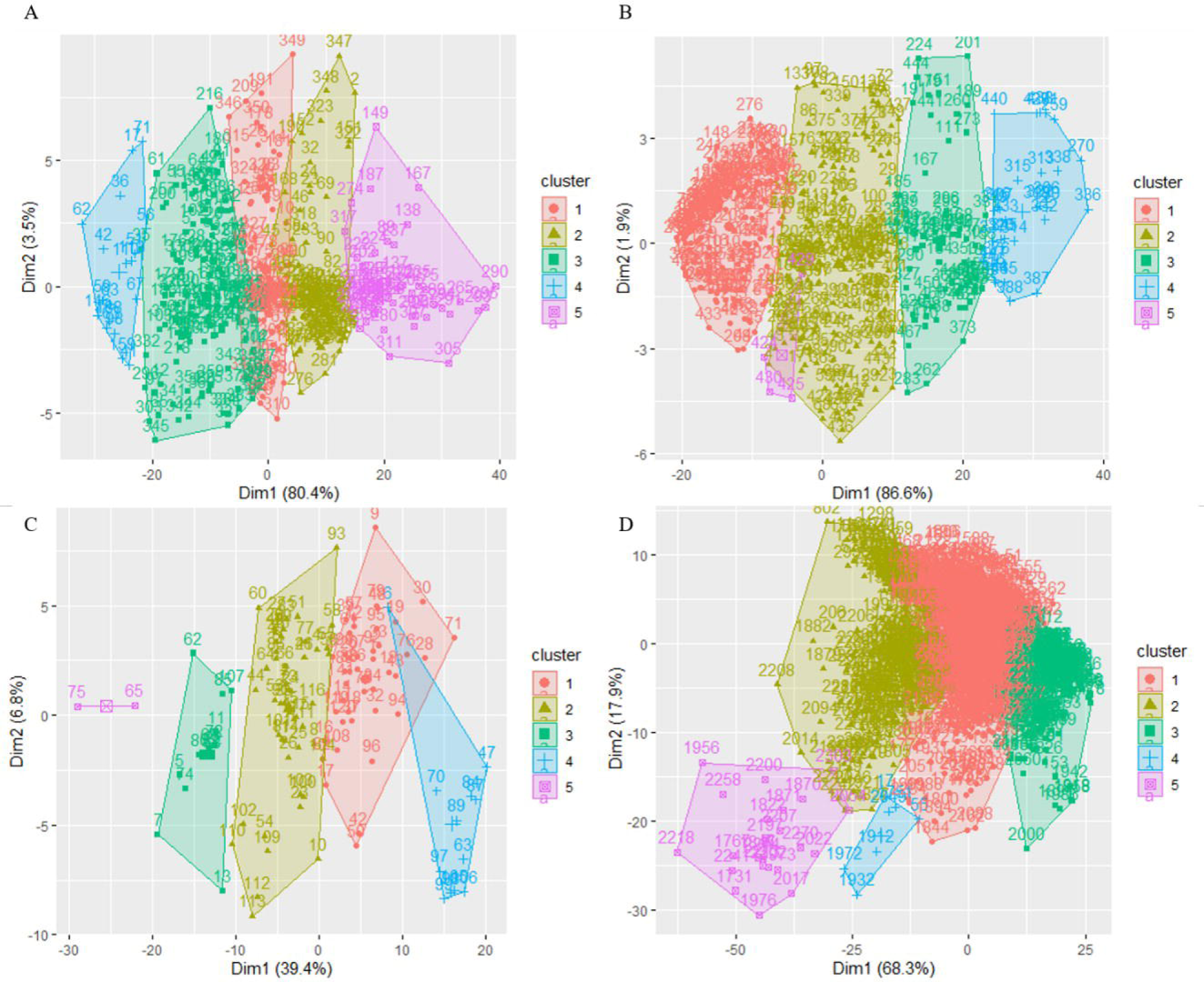
Metabolites are clustered according to similarity at baseline visit. (A-D) Hierarchical clustering of (A) non-polar negative, (B) non-polar positive, (C) polar negative, and (D) polar positive metabolites at baseline. Alt text: Four panel scatterplot figure showing clustering results projected onto two dimensions (Dim1 and Dim2). Each panel displays data points labeled with sample IDs and grouped into five color-coded clusters enclosed by polygon outlines: cluster 1 (filled red circles), cluster 2 (filled yellow triangles), cluster 3 (filled green squares), cluster 4 (blue crosses), and cluster 5 (unfilled purple squares).

Using an unadjusted linear model, we identified 8 clusters correlated with FI score (p<0.05). After adjusting the model for sex and age, 6 clusters remained significantly correlated with FI (p<0.05). Finally, after the model was adjusted for all clinical covariates, the same 6 clusters remained linked with frailty (p<0.05).

Cluster 1 contained 50 metabolites. Over representation analysis found no metabolite classes were significantly enriched within this cluster, however the most common class was glycerophospholipids, followed by prenol lipids. Cluster 2 contained 2 metabolites, which were unable to be identified. Due to lack of identifications and the small size of this cluster, over representation analysis could not be performed. Cluster 3 contained 1758 metabolites and was the largest cluster identified. The most common metabolites significantly enriched within this cluster were carboxylic acids and derivatives, organooxygen compounds, steroids and derivatives, and several species of lipids, with fatty acyls, prenol lipids and sphingolipids the most common. Cluster 4 contained 240 metabolites, and 8 metabolite classes were significantly enriched within the cluster. The largest number of hits were found within carboxylic acid and derivatives and fatty acyl classes. Cluster 5 contained 7 metabolites. Each identified metabolite was mapped to a unique class and as such no classes were significantly enriched within the cluster. Finally, Cluster 6 contained 23 metabolites. Similar to cluster 5, the majority of metabolites belonged to unique classes and were not significantly enriched, however the presence of azolidines was found to be significant (Figure 2A).

**Figure 2.**
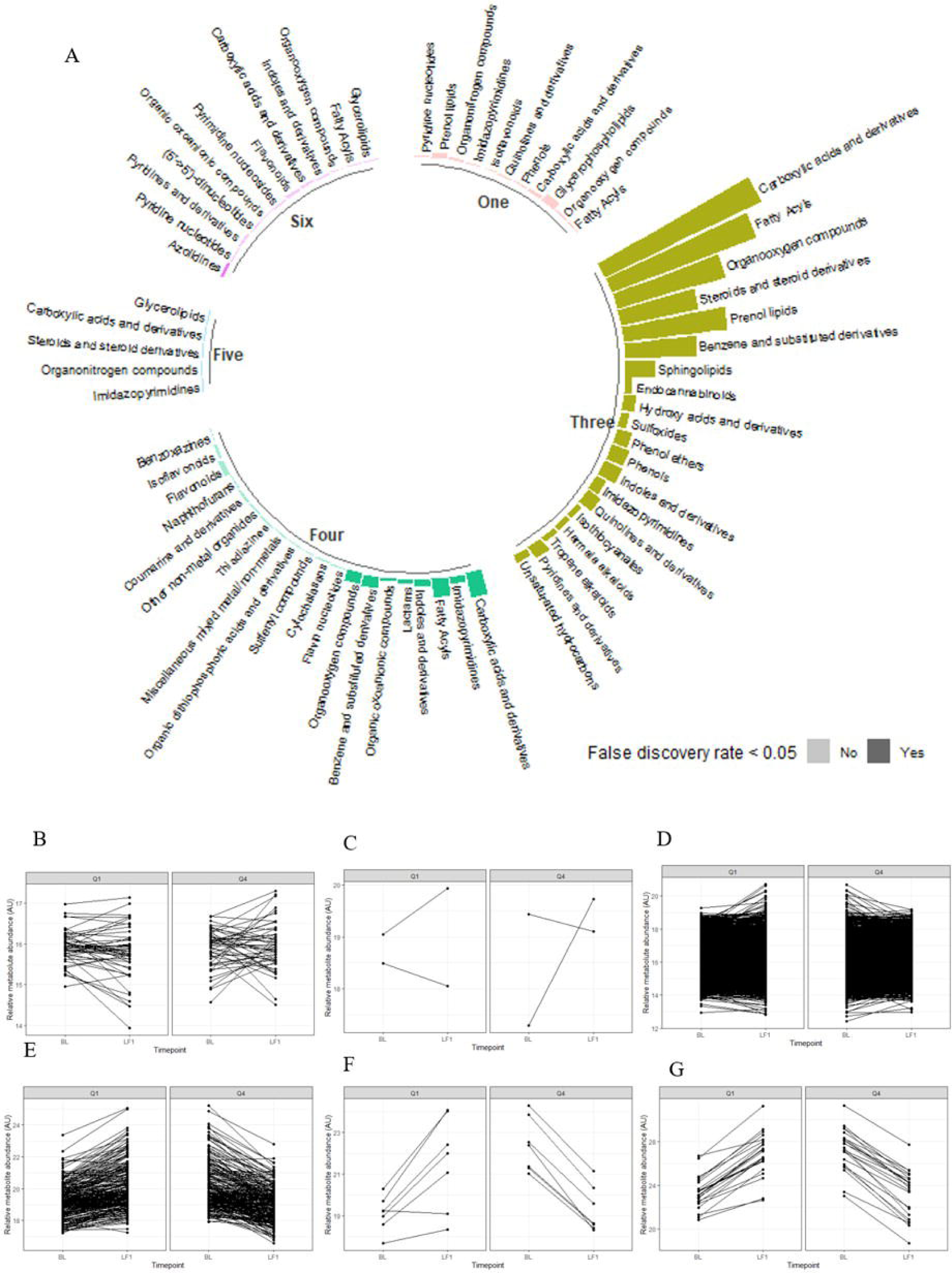
Enrichment of metabolite classes associated with frailty index. (A) Number of hits in identified metabolite classes per cluster at baseline. Significantly enriched classes are highlighted. (B-G) Change in the relative plasma abundance of metabolites (arbitrary units) from baseline to LF1 in (B) cluster 1, (C) cluster 2, (D) cluster 3, (E) cluster 4, (F) cluster 5, and (G) cluster 6. Alt text: Multi-panel figure showing metabolite class enrichment and longitudinal metabolite abundance changes across six clusters. Panel A shows circular bar chart of metabolite class composition for six clusters with shading indicating false discovery rate. Panels B-G show paired line plots of relative metabolite abundance between two time points in quantile 1 and quantile 4 where each line represents an individual metabolite.

There were 40 metabolite classes that were identified in at least 2 clusters. The most common class across these clusters were carboxylic acids and derivatives, which were identified in 5 clusters, however classes such as fatty acyls, glycerolipids, imidazopyramidines, organonitrogen compounds, and organooxygen compounds were also commonly identified, indicating the metabolic networks most likely involved in the aetiology of frailty.

### Plasma metabolite dynamics are dependent on frailty over time

Finally, we examined how plasma abundance of metabolites associated with frailty differed between baseline and LF1 visits in individuals with the lowest (Q1) and highest (Q4) FI scores. We observed distinct patterns of change in metabolite abundance between Q1 and Q4 in each cluster (Figure 2B-G). These patterns were strongest in clusters 2, 4, 5 and 6, where plasma abundance changes in Q1 were the inverse of Q4, indicating that advanced frailty completely alters the composition of the plasma metabolome. Differential abundance analysis found 265 metabolites with p<0.05 (eTable 2 in the Supplement). However, after adjusting for false discovery rate no metabolites remained significantly different, although 65 metabolites across 2 clusters tended towards significance (p*_adj_*<0.1).

## Discussion

Frailty is a known risk factor of dementia and may increase the likelihood of early cognitive decline progressing to dementia. In a cohort of older adults at risk of dementia, we performed global profiling of the plasma metabolome and evaluated the association of plasma metabolites with frailty. We identified 6 clusters of metabolites that were correlated with FI score at baseline. Although each cluster contained unique metabolite identifications, several common metabolite classes were observed within multiple clusters, such as carboxylic acids and derivatives or fatty acyls.

Carboxylic acids and derivatives were the largest class of metabolites in clusters 3 and 4 and were also identified in clusters 1, 5 and 6. Several identified metabolites within these clusters point to a plasma amino acid profile that indicates increased muscle protein breakdown (27). Similar disturbances in amino acid metabolism have previously been linked to frailty in population level association studies (28,29), suggesting loss of muscle mass (30). Exercise interventions have been shown to protect global cognition and executive function in MCI patients (31). Thus, interventions to maintain or increase muscle mass in at risk SCD and MCI patients may slow or prevent progression to dementia, or even stabilise cognitive function in high risk individuals.

Fatty acyls were the second largest class of metabolites in clusters 3 and 4, and were also found in cluster 1, although they were not significantly enriched within this cluster. We identified several octadecanoic acids, which are essential regulators of inflammatory responses, and multiple fatty acids derivatives such as downstream metabolites of linoleic acid or acylcarnitine, which have a variety of physiological roles but notably are essential components in cellular energy metabolismoften derived from dietary fatty acid consumption. Observational studies have previously found that low fatty acid intake, and therefore reduced fatty acid availability, is associated with reduced physical function (32) and a recent metabolomic profiling of the UK Biobank found improved prognosis of frail individuals with high linoleic acid concentrations (28), likely due to its anti-inflammatory properties. Increased acylcarnitine levels have been linked to lower physical ability (33) and functional impairment (34). This increase has been poposed to be indicative of a shift in energy metabolism, aligning with the increased abundance of carboxylic acid derivatives,intermediaries and substrates in glycolysis and the tricarboxylic acid cycle.

Several clusters contained a number of enriched lipid classes including prenol lipids and sphingolipids. This finding is unsurprising, given dysregulated lipid metabolism has been identified as a key factor in both frailty (35) and cognitive impairment (36), with abnormal lipidomic profiles are observed in frail individuals (37) and in the brains of patients with early stage Alzheimer’s disease (38). Notably, sphingolipids have been linked to several forms of dementia (39) and are negatively correlated with muscle mass (40). Sphingolipids have diverse physiological roles and are believed to influence both frailty and dementia through several possible pathways. For instance, the accumulation of sphingolipids can promote neuronal cell death (41) while in muscle, accumulation reduces contractility of muscle fibres, in turn decreasing strength (40). The exact mechanism for the intersection of frailty and dementia through dysregulated sphingolipid metabolism is unclear with further investigation required. The observed association with FI score in SCD and MCI patients highlights the key role of sphingolipids could play as diagnostic markers for individuals most at risk of progressing from early cognitive decline to dementia.

A major metabolite class enriched in cluster 3 was steroids and steroid derivatives. Among the metabolites identified were tetrahydrocorticosterone, a steroid which has previously been identified a biomarker of sarcopenia and is believed to be linked to muscle loss (42), and several downstream metabolites of testosterone. Low testosterone availability has been associated with frailty in both men and women (43), potentially due to inhibited rates of muscle protein synthesis caused by testosterone deficiency in muscle. Additionally, in cluster 5 3alpha,7alpha,12alpha,24(S)-tetrahydroxy-5beta-cholestan-27-al, a bile acid, was identified, although steroids were not enriched within this cluster. Bile acid metabolism is believed to play a critical role in the development of frailty, as bile acids function as signalling molecules which regulate muscle function. Dysregulation of bile acid metabolism disturbs key signalling pathways and contributes to an inflammatory environment (44), promoting muscle wasting.

Organooxygen compounds were identified in clusters 3 and 4 and several metabolites within this class were identified as key components of glucose metabolism central to the tricarboxylic acid cycle and glycolysis pathways. This shift may suggest evidence of metabolic reprogramming, leading to impaired energy metabolism in the progression of frailty. Efficient glucose metabolism and high metabolic activity in the brain are known to protect against cognitive ageing while low brain metabolic activity increases susceptibility to neurodegeneration (45,46). Additionally, a recent multi-omics profiling of transgenic mouse models of Alzheimer’s disease highlighted brain hyperglycosylation and increased glycan biosynthesis, a process tightly linked to glucose metabolism, as a pathological driver of Alzheimer’s disease (47). Furthermore, abnormal glucose uptake in the brain is considered a hallmark of dementia (48), and assessment of ^18^F-FDG uptake by FDG-PET imaging is an established technique used to support differential neurodegenerative diagnoses (49). The detection of plasma metabolites indicative of abnormal glucose metabolism prior to detectable changes in FDG-PET imaging may present an avenue for early diagnosis of at risk individuals. Further research investigating associations between plasma levels of glucose metabolites and glucose uptake in the brain as measured by FDG-PET may also provide greater insight into the precise mechanisms underlying dysregulation of glucose metabolism and the pathophysiology of dementia in at risk patients.

Through longitudinal profiling of the plasma metabolome, we also identified clear differences in the trajectory of plasma metabolite abundance between participants in Q1, who presented with the lowest level of frailty, and participants in Q4, who presented with the highest severity of frailty. We observed a general trend across all clusters indicating that the direction of change in plasma metabolite abundance in Q4 was the inverse of Q1, showing the effect of increased frailty on the plasma metabolome. It should be noted that due to the large size of cluster 3, visually identifying trends within this cluster was not possible due to overlapping metabolite abundances. It is possible that implementing cluster size restrictions may alleviate this issue, however due to the exploratory nature of this study we chose not to impose overly conservative restrictions on unsupervised clustering.

When differential abundance of individual metabolites was assessed, we found no difference in the relative change in abundance over time in Q4 relative to Q1. While tracking of metabolite abundance trajectory showed clear differences, it is possible that rather than attributing this shift to individual metabolites, the frailty-induced shift in metabolism requires multiple metabolites to work in concert. In addition, although no metabolites were significantly changed, there were 65 metabolites with an adjusted p-value of 0.1 or less.

There are several limitations to this study that we acknowledge. Firstly, although frailty is closely linked with low muscle mass and strength, a direct assessment of skeletal muscle strength was not collected at the clinical visit. While BMI, habitual physical activity and daily functional capacity were accounted for the in the calculation of frailty index, the inclusion of measures such as hand grip strength or 6 minute walk test, or a measure of muscle mass through DXA or pQCT may provide a more direct measure of skeletal muscle strength and mass. Ethnicity of participants recruited to this study was largely homogenous, which may limit generalisation of findings. Further validation on a larger representative cohort may be required. Finally, due to the ongoing nature of this study, the number of participants was reduced at each follow up visit. Further analysis should be conducted when sample size at the first and second follow up visits has increased to provide greater statistical power at these timepoints.

To summarise, we have identified a selection of metabolite biomarkers which are closely linked with frailty in older adults most at risk of dementia. These metabolites provide insight into the metabolic pathways and networks disrupted by frailty and may inform possible therapeutic options to mitigate or prevent further progression of frailty and cognitive impairment. We observed wide dysregulation of lipid metabolism in frail individuals. Abnormalities in lipid metabolism have previously been observed at various stages of cognitive decline and appear to be a key component of dementia aetiology. Thus, the initial dysregulation of lipids driven from frailty may accelerate the development of dementia, providing a possible link between increased dementia risk in SCD and MCI patients. Notably, we also identified metabolites linked to utilisation of glucose and energy metabolism, pointing to dysregulation of energy metabolism induced by frailty. This finding aligns with the current understanding of increased glucose uptake in the brain as a hallmark of dementia and presents an appealing avenue for future research and therapeutic options, as efforts to improve or regulate energy metabolism may ameliorate cognitive decline in at risk individuals by reducing the burden of frailty, and detection of glucose metabolites may enable neurodegenerative diagnoses prior to assessment by FDG-PET.

## Data Availability

All data produced in the present study are available upon reasonable request to the authors.

## Acknowledgements

This research was conducted using data provided by the Healthy Brain Ageing Research Program at the Brain and Mind Centre, University of Sydney. We acknowledge the support of the NHMRC Centre of Research Excellence for CogSleep in facilitating this collaboration.

This work was supported by Wellcome Leap’s Dynamic Resilience Program (jointly funded by Temasek Trust) awarded to AP. JCM is supported by the Kennards Hire Foundation Dementia Research Fellowship. This work was facilitated by access to Sydney Mass Spectrometry, a core research facility at the University of Sydney.

We thank Professor Mirana Ramialison, Professor Susan E. Kurrle and Dr Mark Larance for their help, support and advice.

## REFERENCES

1. Global Strategy and Action Plan on Ageing and Health. World Health Organisation; 2017. https://www.who.int/publications/i/item/global-strategy-and-action-plan-on-ageing-and-health.

2. Australian Institute of Health and Welfare. Dementia in Australia. https://www.aihw.gov.au/reports/dementia/dementia-in-aus/contents/summary. Published 13 September 2024. Accessed August 11, 2025.

3. Global Action Plan on the Public Health Response to Dementia 2017-2025. World Health Organisation; 2017.

4. Chrisp TAC, Thomas BD, Goddard WA, Owens A. Dementia timeline: Journeys, delays and decisions on the pathway to an early diagnosis. Dementia. 2011;10(4):555–570.

5. Woods B, Arosio F, Diaz A, et al. Timely diagnosis of dementia? Family carers’ experiences in 5 European countries. Int J Geriatr Psychiatry. 2019;34(1):114–121.

6. Kotagal V, Langa KM, Plassman BL, et al. Factors associated with cognitive evaluations in the United States. Neurology. 2015;84(1):64–71.

7. Jongsiriyanyong S, Limpawattana P. Mild cognitive impairment in clinical practice: A review article. Am J Alzheimers Dis Other Demen. 2018;33(8):500–507.

8. Slot RER, Sikkes SAM, Berkhof J, et al. Subjective cognitive decline and rates of incident Alzheimer’s disease and non-Alzheimer’s disease dementia. Alzheimers Dement. 2019;15(3):465–476.

9. Parfenov VA, Zakharov VV, Kabaeva AR, Vakhnina NV. Subjective cognitive decline as a predictor of future cognitive decline: a systematic review. Dement Neuropsychol. 2020;14(3):248–257.

10. Livingston G, Huntley J, Liu KY, et al. Dementia prevention, intervention, and care: 2024 report of the Lancet standing Commission. Lancet. 2024;404(10452):572–628.

11. Fried LP, Tangen CM, Walston J, et al. Frailty in older adults: evidence for a phenotype. J Gerontol A Biol Sci Med Sci. 2001;56(3):M146–56.

12. Rockwood K, Song X, MacKnight C, et al. A global clinical measure of fitness and frailty in elderly people. CMAJ. 2005;173(5):489–495.

13. Cho Y, Huang J, Liu S, Lukkahatai N, Gill J, Li J. Evaluating physical frailty as a marker of cognitive health in older adults without dementia. J Geriatr Psychiatry Neurol. January 2026:8919887261421225.

14. Cermelli A, Crisafi A, Chiarandon AM, et al. Exploring frailty status and blood biomarkers: A multidimensional approach to Alzheimer’s diagnosis. Geriatrics (Basel). 2025;10(5):133.

15. Cullen NC, Leuzy A, Janelidze S, et al. Plasma biomarkers of Alzheimer’s disease improve prediction of cognitive decline in cognitively unimpaired elderly populations. Nat Commun. 2021;12(1):3555.

16. Grande G, Valletta M, Rizzuto D, et al. Blood-based biomarkers of Alzheimer’s disease and incident dementia in the community. Nat Med. 2025;31(6):2027–2035.

17. Corso G, Cristofano A, Sapere N, et al. Serum amino acid profiles in normal subjects and in patients with or at risk of Alzheimer dementia. Dement Geriatr Cogn Dis Extra. 2017;7(1):143–159.

18. Milos T, Rojo D, Nedic Erjavec G, et al. Metabolic profiling of Alzheimer’s disease: Untargeted metabolomics analysis of plasma samples. Prog Neuropsychopharmacol Biol Psychiatry. 2023;127(110830):110830.

19. Searle SD, Mitnitski A, Gahbauer EA, Gill TM, Rockwood K. A standard procedure for creating a frailty index. BMC Geriatr. 2008;8(1):24.

20. Sostare J, Di Guida R, Kirwan J, et al. Comparison of modified Matyash method to conventional solvent systems for polar metabolite and lipid extractions. Anal Chim Acta. 2018;1037:301–315.

21. R Core Team. R: A Language and Environment for Statistical Computing. 2021. https://www.R-project.org/.

22. Smith CA, Want EJ, O’Maille G, Abagyan R, Siuzdak G. XCMS: processing mass spectrometry data for metabolite profiling using nonlinear peak alignment, matching, and identification. Anal Chem. 2006;78(3):779–787.

23. Broadhurst D, Goodacre R, Reinke SN, et al. Guidelines and considerations for the use of system suitability and quality control samples in mass spectrometry assays applied in untargeted clinical metabolomic studies. Metabolomics. 2018;14(6):72.

24. Shen X, Yan H, Wang C, Gao P, Johnson CH, Snyder MP. TidyMass an object-oriented reproducible analysis framework for LC-MS data. Nat Commun. 2022;13(1):4365.

25. Pang Z, Chong J, Zhou G, et al. MetaboAnalyst 5.0: narrowing the gap between raw spectra and functional insights. Nucleic Acids Res. 2021;49(W1):W388–W396.

26. Ritchie ME, Phipson B, Wu D, et al. limma powers differential expression analyses for RNA-sequencing and microarray studies. Nucleic Acids Res. 2015;43(7):e47.

27. Calvani R, Picca A, Marini F, et al. A distinct pattern of circulating amino acids characterizes older persons with Physical frailty and sarcopenia: Results from the BIOSPHERE study. Nutrients. 2018;10(11):1691.

28. Xiao L, Deng Q, Wang J, et al. Metabolomic characterization of frailty identifies subtype-specific management strategies. NPJ Digit Med. 2025;8(1):746.

29. Shekarchian A, Bandarian F, Hadizadeh A, et al. Exploring the metabolomics profile of frailty- a systematic review. J Diabetes Metab Disord. 2024;23(1):289–303.

30. Dai M, Lin T, Yue J, Dai L. Signatures and clinical significance of amino acid flux in sarcopenia: A systematic review and meta-analysis. Front Endocrinol (Lausanne). 2021;12:725518.

31. Huang X, Zhao X, Li B, et al. Comparative efficacy of various exercise interventions on cognitive function in patients with mild cognitive impairment or dementia: A systematic review and network meta-analysis. J Sport Health Sci. 2022;11(2):212–223.

32. Kim D, Won CW, Park Y. Association between erythrocyte levels of n-3 polyunsaturated fatty acids and risk of frailty in community-dwelling older adults: The Korean Frailty and Aging Cohort Study. J Gerontol A Biol Sci Med Sci. 2021;76(3):499–504.

33. Lum H, Sloane R, Huffman KM, et al. Plasma acylcarnitines are associated with physical performance in elderly men. J Gerontol A Biol Sci Med Sci. 2011;66(5):548–553.

34. Caballero FF, Struijk EA, Lana A, Buño A, Rodríguez-Artalejo F, Lopez-Garcia E. Plasma acylcarnitines and risk of lower-extremity functional impairment in older adults: a nested case-control study. Sci Rep. 2021;11(1):3350.

35. Mak JKL, Kananen L, Qin C, et al. Unraveling the metabolic underpinnings of frailty using multicohort observational and Mendelian randomization analyses. Aging Cell. 2023;22(8):e13868.

36. Anand SS, Friedrich MG, Lee DS, et al. Evaluation of adiposity and cognitive function in adults. JAMA Netw Open. 2022;5(2):e2146324.

37. Ramírez-Vélez R, Martínez-Velilla N, Correa-Rodríguez M, et al. Lipidomic signatures from physically frail and robust older adults at hospital admission. GeroScience. 2022;44(3):1677–1688.

38. Naudí A, Cabré R, Jové M, et al. Lipidomics of human brain aging and Alzheimer’s disease pathology. Int Rev Neurobiol. 2015;122:133–189.

39. Moseholm KF, Cronjé HT, Koch M, et al. Circulating sphingolipids in relation to cognitive decline and incident dementia: The Cardiovascular Health Study. Alzheimers Dement (Amst). 2024;16(3):e12623.

40. Laurila P-P, Wohlwend M, Imamura de Lima T, et al. Sphingolipids accumulate in aged muscle, and their reduction counteracts sarcopenia. Nat Aging. 2022;2(12):1159–1175.

41. Satoi H, Tomimoto H, Ohtani R, et al. Astroglial expression of ceramide in Alzheimer’s disease brains: a role during neuronal apoptosis. Neuroscience. 2005;130(3):657–666.

42. Wu H, Gao H, Guo L, et al. Creatinine, leucine, and tetrahydrocorticosterone emerge as potential biomarkers for the diagnosis of sarcopenic osteoarthritis in middle-aged and elderly individuals: a cross-sectional exploratory study. Front Mol Biosci. 2026;13:1743095.

43. Wu I-C, Lin X-Z, Liu P-F, Tsai W-L, Shiesh S-C. Low serum testosterone and frailty in older men and women. Maturitas. 2010;67(4):348–352.

44. Sun J, Zhang S, Jin L, Huang W. Bile acid signaling, metabolism, and aging. Liver Res. February 2026. doi:10.1016/j.livres.2026.02.002.

45. Kapogiannis D, Mattson MP. Disrupted energy metabolism and neuronal circuit dysfunction in cognitive impairment and Alzheimer’s disease. Lancet Neurol. 2011;10(2):187–198.

46. Yin F, Sancheti H, Patil I, Cadenas E. Energy metabolism and inflammation in brain aging and Alzheimer’s disease. Free Radic Biol Med. 2016;100:108–122.

47. Hawkinson TR, Liu Z, Ribas RA, et al. Hyperglycosylation is a metabolic driver of Alzheimer’s disease. Nat Metab. 2026;8(6):1410–1425.

48. Garrett LR, Niccoli T. Frontotemporal dementia and glucose metabolism. Front Neurosci. 2022;16:812222.

49. Minoshima S, Cross D, Thientunyakit T, Foster NL, Drzezga A. 18F-FDG PET imaging in neurodegenerative dementing disorders: Insights into subtype classification, emerging disease categories, and mixed dementia with copathologies. J Nucl Med. 2022;63(Suppl 1):2S–12S.

