## Supplementary Table 1 for "Plasma metabolomic profiling of frailty in older adults at risk of dementia"

**eTable 1. Full list of health deficits included in the Frailty Index**

| **No.** | **Variable** | **Coding with assigned value for Frailty Index** |
| --- | --- | --- |
|  | **Chronic diseases / medical history** |  |
| 1 | Hypercholesterolaemia | “Yes” = 1; “No” = 0 |
| 2 | Stroke | “Yes” = 1; “No” = 0 |
| 3 | Thyroid disease | “Yes” = 1; “No” = 0 |
| 4 | Chronic pain | “Yes” = 1; “No” = 0 |
| 5 | Cancer or tumour | “Yes” = 1; “No” = 0 |
| 6 | Hypertension | “Yes” = 1; “No” = 0 |
| 7 | Diabetes | “Yes” = 1; “No” = 0 |
| 8 | Cognitive/sleep-related condition | “Yes” = 1; “No” = 0 |
| 9 | Epilepsy or seizures | “Yes” = 1; “No” = 0 |
| 10 | Other neurological disorder | “Yes” = 1; “No” = 0 |
| 11 | Other serious medical illness | “Yes” = 1; “No” = 0 |
| 12 | Severe infection | “Yes” = 1; “No” = 0 |
|  | **Anthropometry / weight-related deficits** |  |
| 13 | Body mass index | Deficit coded according to BMI category |
| 14 | Waist circumference | Elevated waist circumference = 1; not elevated = 0 |
|  | **Comorbidity burden / organ system morbidity** |  |
| 15 | Cardiac morbidity | Deficit coded from CIRS heart rating |
| 16 | Vascular morbidity | Deficit coded from CIRS vascular rating |
| 17 | Haematopoietic morbidity | Deficit coded from CIRS haematopoietic rating |
| 18 | Respiratory morbidity | Deficit coded from CIRS respiratory rating |
| 19 | Eye, ear, nose, throat, or larynx morbidity | Deficit coded from CIRS eyes/ears/nose/throat/larynx rating |
| 20 | Upper gastrointestinal morbidity | Deficit coded from CIRS upper gastrointestinal rating |
| 21 | Lower gastrointestinal morbidity | Deficit coded from CIRS lower gastrointestinal rating |
| 22 | Liver morbidity | Deficit coded from CIRS liver rating |
| 23 | Renal morbidity | Deficit coded from CIRS renal rating |
| 24 | Genitourinary morbidity | Deficit coded from CIRS genitourinary rating |
| 25 | Musculoskeletal morbidity | Deficit coded from CIRS musculoskeletal rating |
| 26 | Neurological morbidity | Deficit coded from CIRS neurological rating |
|  | **Mood, wellbeing, and lifestyle factors** |  |
| 27 | Depression or elevated depressive symptoms | Deficit coded according to depression measure/category |
| 28 | Low physical activity / exercise deficit | Insufficient exercise = 1; sufficient exercise = 0 |
| 29 | Reduced wellbeing | Deficit coded according to wellbeing measure/category |
|  | **Medication burden** |  |
| 30 | Polypharmacy | Polypharmacy present = 1; absent = 0 |

**Note.** FI = Frailty Index; CIRS = Cumulative Illness Rating Scale. Individual deficits were coded from 0 to 1, where higher scores indicate greater deficit severity. The FI was calculated as the sum of deficits divided by the number of non-missing deficits available for each participant.
