## Supplementary Table 2 for "Plasma metabolomic profiling of frailty in older adults at risk of dementia"

**eTable 2. Full list of included metabolites**

Table displaying metabolite study identifier (study ID), common metabolite name and related HMDB identifier, log fold change (logFC), unadjusted p value, p value following adjustment for false discovery rate using the Benjamini-Hochberg method, and cluster where metabolite was detected of all metabolites with an unadjusted p-value<0.05 following differential abundance analysis. p_adj_<0.1 are highlighted in bold.

| **Metabolite study ID** | **Metabolite name** | **HMDB ID** | **logFC** | **p-value** | **Adjusted p-value** | **Cluster** |
| --- | --- | --- | --- | --- | --- | --- |
| METAB212 | NA | NA | 2.880 | 0.0273 | **0.0545** | 2 |
| METAB8853 | NA | NA | -4.341 | 0.0010 | **0.0691** | 4 |
| METAB8205 | NA | NA | -3.210 | 0.0012 | **0.0691** | 4 |
| METAB8242 | 5-(3',4',5'-Trihydroxyphenyl)-gamma-valerolactone-3'-O-sulphate | HMDB0059985 | -3.142 | 0.0014 | **0.0691** | 4 |
| METAB7991 | NA | NA | -2.961 | 0.0017 | **0.0691** | 4 |
| METAB7443 | {1-[5-hydroxy-8-(2-hydroxypropan-2-yl)-2-oxo-2H,8H,9H-furo[2,3-h]chromen-4-yl]propoxy}sulfonic acid | HMDB0130139 | -2.813 | 0.0018 | **0.0691** | 4 |
| METAB8504 | Chloramphenicol | HMDB0014589 | -3.212 | 0.0022 | **0.0691** | 4 |
| METAB8708 | NA | NA | -3.104 | 0.0027 | **0.0691** | 4 |
| METAB7708 | (S)-Spirobrassinin | HMDB0035974 | -2.160 | 0.0036 | **0.0691** | 4 |
| METAB7962 | NA | NA | -2.980 | 0.0036 | **0.0691** | 4 |
| METAB8224 | 6-{[3-(5,7-dihydroxy-2,2-dimethyl-3,4-dihydro-2H-1-benzopyran-6-yl)-4-oxo-4H-chromen-7-yl]oxy}-3,4,5-trihydroxyoxane-2-carboxylic acid | HMDB0125997 | -2.777 | 0.0042 | **0.0691** | 4 |
| METAB7473 | Glucocleomin | HMDB0034045 | -3.050 | 0.0042 | **0.0691** | 4 |
| METAB8210 | Isomelitric acid A | HMDB0039523 | -3.211 | 0.0045 | **0.0691** | 4 |
| METAB8219 | NA | NA | -2.656 | 0.0048 | **0.0691** | 4 |
| METAB9080 | NA | NA | -4.427 | 0.0049 | **0.0691** | 4 |
| METAB8943 | erythro-6,8-Pentacosanediol | HMDB0041075 | -4.375 | 0.0050 | **0.0691** | 4 |
| METAB6702 | 2-(Methylthio)-3H-phenoxazin-3-one | HMDB0035996 | -2.680 | 0.0050 | **0.0691** | 4 |
| METAB8767 | Triazolam | HMDB0015034 | -3.339 | 0.0054 | **0.0691** | 4 |
| METAB6236 | 6-Methylmercaptopurine | HMDB0060412 | -2.788 | 0.0055 | **0.0691** | 4 |
| METAB694 | NA | NA | 3.051 | 0.0058 | **0.0691** | 4 |
| METAB9188 | PC(24:0/24:0) | HMDB0008782 | -4.637 | 0.0062 | **0.0691** | 4 |
| METAB7419 | 3-Methylpentyl glucosinolate | HMDB0036355 | -2.512 | 0.0062 | **0.0691** | 4 |
| METAB9119 | Delphinidin 3-lathyroside 5-glucoside | HMDB0037090 | -3.772 | 0.0063 | **0.0691** | 4 |
| METAB5701 | alpha-Fluoro-beta-alanine | HMDB0060434 | -4.121 | 0.0071 | **0.0710** | 4 |
| METAB6825 | MG(0:0/16:0/0:0) | HMDB0011533 | -3.919 | 0.0072 | **0.0710** | 4 |
| METAB7460 | dCDP | HMDB0001245 | -2.128 | 0.0074 | **0.0710** | 4 |
| METAB8317 | Clofarabine | HMDB0014769 | -5.052 | 0.0091 | **0.0777** | 4 |
| METAB7984 | 3,4,5-trihydroxy-6-{[3,6,7-trihydroxy-2-(4-hydroxy-3-methoxyphenyl)-4-oxo-4H-chromen-5-yl]oxy}oxane-2-carboxylic acid | HMDB0128769 | -3.006 | 0.0091 | **0.0777** | 4 |
| METAB9111 | Heterophylliin E | HMDB0039268 | -3.473 | 0.0092 | **0.0777** | 4 |
| METAB9126 | Shoyuflavone C | HMDB0034590 | -4.163 | 0.0098 | **0.0777** | 4 |
| METAB8805 | NA | NA | -3.915 | 0.0098 | **0.0777** | 4 |
| METAB8589 | 3-(3,7-dihydroxy-5-sulfino-3,4-dihydro-2H-1-benzopyran-2-yl)benzen-1-olate | HMDB0127799 | -6.972 | 0.0102 | **0.0777** | 4 |
| METAB9132 | NA | NA | -3.702 | 0.0104 | **0.0777** | 4 |
| METAB6567 | NA | NA | -3.192 | 0.0110 | **0.0798** | 4 |
| METAB2442 | NA | NA | -0.885 | 0.0119 | **0.0837** | 4 |
| METAB9097 | 3,4,5-trihydroxy-6-{[7,8,8,13,22-pentahydroxy-19-(hydroxymethyl)-3,6,16-trioxo-21-(3,4,5-trihydroxybenzoyloxy)-2,17,20,23-tetraoxapentacyclotricosa-4,10,12,14-tetraen-12-yl]oxy}oxane-2-carboxylic acid | HMDB0126672 | -4.832 | 0.0136 | **0.0932** | 4 |
| METAB7963 | 2-amino-4-({1-[(carboxymethyl)-C-hydroxycarbonimidoyl]-2-{[1,3-dihydroxy-1-(2-hydroxyphenyl)propan-2-yl]sulfanyl}ethyl}-C-hydroxycarbonimidoyl)butanoic acid | HMDB0134078 | -2.169 | 0.0147 | **0.0932** | 4 |
| METAB7670 | NA | NA | -6.492 | 0.0152 | **0.0932** | 4 |
| METAB9153 | NA | NA | -4.658 | 0.0152 | **0.0932** | 4 |
| METAB7215 | Phosphoguanidinoacetate | HMDB0003705 | -2.756 | 0.0164 | **0.0932** | 4 |
| METAB6029 | Butyric acid | HMDB0000039 | -3.273 | 0.0170 | **0.0932** | 4 |
| METAB8055 | NA | NA | -3.435 | 0.0172 | **0.0932** | 4 |
| METAB8379 | 5alpha-Ethoxy-6beta-hydroxy-5,6-dihydrophysalin B | HMDB0029626 | -3.096 | 0.0173 | **0.0932** | 4 |
| METAB7569 | NA | NA | -2.501 | 0.0174 | **0.0932** | 4 |
| METAB7980 | NA | NA | -3.091 | 0.0175 | **0.0932** | 4 |
| METAB9143 | 6-Hydroxy flavin adenine dinucleotide | HMDB0011612 | -6.271 | 0.0178 | **0.0932** | 4 |
| METAB7958 | 3,4,5-trihydroxy-6-{[3,6,7-trihydroxy-2-(4-hydroxy-3-methoxyphenyl)-4-oxo-3,4-dihydro-2H-1-benzopyran-5-yl]oxy}oxane-2-carboxylic acid | HMDB0128791 | -3.181 | 0.0179 | **0.0932** | 4 |
| METAB817 | L-2-Amino-4-methylenepentanedioic acid | HMDB0029433 | 1.072 | 0.0185 | **0.0937** | 4 |
| METAB7312 | NA | NA | -2.444 | 0.0187 | **0.0937** | 4 |
| METAB7197 | Acenocoumarol | HMDB0015487 | -3.366 | 0.0198 | **0.0938** | 4 |
| METAB8037 | NA | NA | -4.350 | 0.0200 | **0.0938** | 4 |
| METAB5815 | 2-[(Methylthio)methyl]-2-butenal | HMDB0031370 | -2.393 | 0.0201 | **0.0938** | 4 |
| METAB7672 | 6-(3,4-dihydroxy-6-methyl-5-oxooxan-2-yl)-5,7-dihydroxy-2-(3-methoxyphenyl)-4H-chromen-4-one | HMDB0127330 | -2.339 | 0.0213 | **0.0938** | 4 |
| METAB8473 | TG(12:0/12:0/12:0) | HMDB0011188 | -2.652 | 0.0214 | **0.0938** | 4 |
| METAB3364 | NA | NA | -1.040 | 0.0214 | **0.0938** | 4 |
| METAB8188 | NA | NA | -2.873 | 0.0217 | **0.0938** | 4 |
| METAB8726 | NA | NA | -4.560 | 0.0219 | **0.0938** | 4 |
| METAB7829 | 3,4,5-trihydroxy-6-[3-(3,5,7-trihydroxy-5H-chromen-2-yl)phenoxy]oxane-2-carboxylic acid | HMDB0127942 | -1.642 | 0.0223 | **0.0940** | 4 |
| METAB7320 | NA | NA | -2.507 | 0.0233 | **0.0963** | 4 |
| METAB6291 | N-Acetyl-L-aspartic acid | HMDB0000812 | -3.602 | 0.0237 | **0.0964** | 4 |
| METAB9046 | Metrizamide | HMDB0015518 | -3.260 | 0.0247 | **0.0986** | 4 |
| METAB7882 | LysoPC(0:0/16:0) | HMDB0240262 | -2.318 | 0.0261 | **0.0999** | 4 |
| METAB7989 | 3,4,5-trihydroxy-6-{[3,6,7-trihydroxy-2-(4-hydroxy-3-methoxyphenyl)-4-oxo-4H-chromen-5-yl]oxy}oxane-2-carboxylic acid | HMDB0128769 | -4.931 | 0.0263 | **0.0999** | 4 |
| METAB7872 | Didodecyl thiobispropanoate | HMDB0040172 | -3.323 | 0.0263 | **0.0999** | 4 |
| METAB7033 | NA | NA | -2.318 | 0.0267 | **0.0999** | 4 |
| METAB7688 | Halosulfuron-methyl | HMDB0034859 | -3.116 | 0.0277 | 0.1007 | 4 |
| METAB6409 | Distichonic acid A | HMDB0038752 | -6.431 | 0.0277 | 0.1007 | 4 |
| METAB7566 | NA | NA | -2.475 | 0.0293 | 0.1033 | 4 |
| METAB8865 | C.I. Food Black 2 | HMDB0033393 | -5.193 | 0.0294 | 0.1033 | 4 |
| METAB8008 | 3,4,5-trihydroxy-6-{[3,5,7-trihydroxy-4-oxo-2-(3,4,5-trihydroxyphenyl)-3,4-dihydro-2H-1-benzopyran-6-yl]oxy}oxane-2-carboxylic acid | HMDB0125289 | -4.143 | 0.0300 | 0.1033 | 4 |
| METAB7043 | Erythrityl Tetranitrate | HMDB0015551 | -5.518 | 0.0304 | 0.1033 | 4 |
| METAB7387 | Cefdinir | HMDB0014675 | -2.056 | 0.0306 | 0.1033 | 4 |
| METAB7842 | LysoPE(0:0/22:6(4Z,7Z,10Z,13Z,16Z,19Z)) | HMDB0011496 | -3.122 | 0.0325 | 0.1075 | 4 |
| METAB8013 | Tetrahydroaldosterone-3-glucuronide | HMDB0010357 | -4.973 | 0.0327 | 0.1075 | 4 |
| METAB8757 | NA | NA | -2.542 | 0.0331 | 0.1075 | 4 |
| METAB8957 | 5-(acetyloxy)-4,14-dihydroxy-9-oxo-8,17-dioxatetracycloheptadeca-1(10),2,4,6,11,13,15-heptaen-13-yl acetate | HMDB0128412 | -1.664 | 0.0342 | 0.1094 | 4 |
| METAB6570 | Uridine | HMDB0000296 | -10.397 | 0.0135 | 0.1105 | 6 |
| METAB7681 | 3,4,5-trihydroxy-6-{3-[2-(3-hydroxyphenyl)ethyl]-5-methoxyphenoxy}oxane-2-carboxylic acid | HMDB0129923 | -9.879 | 0.0142 | 0.1105 | 6 |
| METAB8884 | NA | NA | -10.147 | 0.0227 | 0.1105 | 6 |
| METAB7697 | Nitrofurantoin | HMDB0014836 | -8.127 | 0.0239 | 0.1105 | 6 |
| METAB7879 | Nb-Lignoceroyltryptamine | HMDB0040819 | -6.350 | 0.0363 | 0.1105 | 6 |
| METAB6722 | Chlorpheniramine | HMDB0001944 | -7.418 | 0.0390 | 0.1105 | 6 |
| METAB8696 | Nicotinic acid adenine dinucleotide | HMDB0001179 | -8.610 | 0.0396 | 0.1105 | 6 |
| METAB6728 | N-Benzoylaspartic acid | HMDB0034251 | -8.027 | 0.0416 | 0.1105 | 6 |
| METAB5751 | Betaine | HMDB0000043 | -9.015 | 0.0459 | 0.1105 | 6 |
| METAB9124 | Glycerol 1,3-dihexadecanoate 2-(9Z-octadecenoate) | HMDB0030969 | -2.823 | 0.0358 | 0.1126 | 4 |
| METAB7711 | (S)-Spirobrassinin | HMDB0035974 | -3.038 | 0.0361 | 0.1126 | 4 |
| METAB6665 | Pentadecanoic acid | HMDB0000826 | -2.637 | 0.0375 | 0.1150 | 4 |
| METAB7457 | Ceftizoxime | HMDB0015427 | -2.113 | 0.0379 | 0.1150 | 4 |
| METAB5997 | NA | NA | -3.977 | 0.0386 | 0.1150 | 4 |
| METAB7682 | Amiloride | HMDB0014732 | -1.808 | 0.0389 | 0.1150 | 4 |
| METAB7578 | NA | NA | -4.266 | 0.0393 | 0.1150 | 4 |
| METAB7280 | Diethyldithiophosphate | HMDB0061736 | -2.681 | 0.0408 | 0.1152 | 4 |
| METAB8801 | NA | NA | -3.070 | 0.0413 | 0.1152 | 4 |
| METAB6503 | Acetaminophen cystein | HMDB0060559 | -2.205 | 0.0414 | 0.1152 | 4 |
| METAB7838 | 2,3,4-Trimethyltriacontane | HMDB0029986 | -5.637 | 0.0417 | 0.1152 | 4 |
| METAB7480 | NA | NA | -3.741 | 0.0417 | 0.1152 | 4 |
| METAB9023 | TG(14:0/15:0/o-18:0) | HMDB0042120 | -2.578 | 0.0432 | 0.1176 | 4 |
| METAB7329 | 3-Hydroxynonyl acetate | HMDB0032443 | -2.560 | 0.0436 | 0.1176 | 4 |
| METAB7247 | NA | NA | -7.864 | 0.0353 | 0.1203 | 5 |
| METAB9107 | PS(15:0/24:0) | HMDB0112340 | -3.965 | 0.0459 | 0.1223 | 4 |
| METAB3678 | Nb-Arachidoyltryptamine | HMDB0040817 | 1.015 | 0.0471 | 0.1242 | 4 |
| METAB6069 | Benzphetamine | HMDB0015003 | -6.065 | 0.0490 | 0.1265 | 4 |
| METAB6742 | 1',2'-Dihydro-1,1'-dimethyl-2'-oxo-4,4'-bipyridinium(1+) | HMDB0035170 | -4.460 | 0.0490 | 0.1265 | 4 |
| METAB25 | 3-Methylxanthine | HMDB0001886 | 3.054 | 0.0118 | 0.4493 | 1 |
| METAB176 | Tuftsin | HMDB0005770 | 1.560 | 0.0246 | 0.4493 | 1 |
| METAB88 | Dibutyl disulfide | HMDB0029569 | -2.279 | 0.0270 | 0.4493 | 1 |
| METAB6301 | 1-(4-methoxyphenyl)-4-methylpenta-1,4-dien-3-one | HMDB0132938 | -2.434 | 0.0004 | 0.5047 | 3 |
| METAB8756 | NA | NA | -1.405 | 0.0006 | 0.5047 | 3 |
| METAB164 | 3-Methoxy-4-hydroxyphenylethyleneglycol sulfate | HMDB0000559 | 2.202 | 0.0009 | 0.5047 | 3 |
| METAB22 | Phosphorylcholine | HMDB0001565 | 2.614 | 0.0414 | 0.5174 | 1 |
| METAB1998 | Ubiquinone-2 | HMDB0006709 | -1.170 | 0.0014 | 0.5254 | 3 |
| METAB298 | Didodecyl thiobispropanoate | HMDB0040172 | -0.844 | 0.0018 | 0.5254 | 3 |
| METAB889 | 5-(2-Methylpropyl)tetrahydro-2-oxo-3-furancarboxylic acid | HMDB0030988 | -0.505 | 0.0032 | 0.5254 | 3 |
| METAB6317 | 4-(Dimethylamino)azobenzene | HMDB0032141 | -2.982 | 0.0036 | 0.5254 | 3 |
| METAB161 | Cardanolmonoene | HMDB0030482 | 1.647 | 0.0040 | 0.5254 | 3 |
| METAB5610 | NA | NA | -0.611 | 0.0045 | 0.5254 | 3 |
| METAB469 | PA(18:4(6Z,9Z,12Z,15Z)/22:6(4Z,7Z,10Z,13Z,16Z,19Z)) | HMDB0115051 | 1.560 | 0.0046 | 0.5254 | 3 |
| METAB5452 | PE-NMe2(22:0/24:1(15Z)) | HMDB0114436 | -0.678 | 0.0065 | 0.5254 | 3 |
| METAB2459 | Indoleacetyl glutamine | HMDB0013240 | -1.304 | 0.0066 | 0.5254 | 3 |
| METAB3628 | 16-Acetylpriverogenin A | HMDB0034529 | -0.622 | 0.0066 | 0.5254 | 3 |
| METAB1908 | Vinylacetylglycine | HMDB0000894 | -1.436 | 0.0071 | 0.5254 | 3 |
| METAB3677 | Tsugaric acid A | HMDB0032022 | -0.829 | 0.0073 | 0.5254 | 3 |
| METAB6700 | Voglibose | HMDB0015598 | -2.288 | 0.0076 | 0.5254 | 3 |
| METAB7232 | gamma-Crocetin | HMDB0035099 | -2.285 | 0.0087 | 0.5254 | 3 |
| METAB541 | (S)-3-Mercaptohexyl pentanoate | HMDB0038945 | -1.086 | 0.0087 | 0.5254 | 3 |
| METAB8699 | NA | NA | -5.539 | 0.0089 | 0.5254 | 3 |
| METAB1835 | NA | NA | -0.707 | 0.0091 | 0.5254 | 3 |
| METAB5074 | NA | NA | -0.635 | 0.0095 | 0.5254 | 3 |
| METAB7406 | Lactapiperanol D | HMDB0033631 | -2.222 | 0.0100 | 0.5254 | 3 |
| METAB8335 | N-Acetylaspartylglutamic acid | HMDB0001067 | -1.202 | 0.0100 | 0.5254 | 3 |
| METAB2558 | 5a-Cholesta-8,24-dien-3-one | HMDB0001093 | -0.696 | 0.0100 | 0.5254 | 3 |
| METAB5494 | NA | NA | -0.516 | 0.0101 | 0.5254 | 3 |
| METAB7786 | NA | NA | -3.770 | 0.0102 | 0.5254 | 3 |
| METAB8773 | Dipivefrin | HMDB0014592 | -1.297 | 0.0105 | 0.5254 | 3 |
| METAB8918 | N-Desmethyleletriptan | HMDB0013919 | -1.239 | 0.0108 | 0.5254 | 3 |
| METAB558 | Humulinic acid A | HMDB0030104 | -2.612 | 0.0110 | 0.5254 | 3 |
| METAB281 | LysoPE(0:0/18:1(11Z)) | HMDB0011475 | 1.046 | 0.0114 | 0.5254 | 3 |
| METAB5146 | NA | NA | -0.739 | 0.0115 | 0.5254 | 3 |
| METAB7945 | 3-(3,5-dihydroxyphenyl)-2-(4-hydroxyphenyl)-4-[(E)-2-(4-hydroxyphenyl)ethenyl]-2,3-dihydro-1-benzofuran-5,6-diol | HMDB0129148 | -2.602 | 0.0116 | 0.5254 | 3 |
| METAB8115 | NA | NA | -1.027 | 0.0122 | 0.5254 | 3 |
| METAB6053 | Capryloylglycine | HMDB0000832 | -1.363 | 0.0124 | 0.5254 | 3 |
| METAB3667 | Carpaine | HMDB0030272 | -0.692 | 0.0126 | 0.5254 | 3 |
| METAB5740 | Allantoic acid | HMDB0001209 | -2.003 | 0.0135 | 0.5254 | 3 |
| METAB5068 | NA | NA | -1.240 | 0.0138 | 0.5254 | 3 |
| METAB5697 | 2-(methylamino)ethan-1-ol | HMDB0142449 | -2.434 | 0.0139 | 0.5254 | 3 |
| METAB444 | AS 1-5 | HMDB0032843 | 1.185 | 0.0145 | 0.5254 | 3 |
| METAB8914 | Physagulin E | HMDB0039693 | -1.271 | 0.0146 | 0.5254 | 3 |
| METAB6578 | 3-Methylglutarylcarnitine | HMDB0000552 | -1.497 | 0.0151 | 0.5254 | 3 |
| METAB446 | Tetrahydrocorticosterone | HMDB0000268 | 1.142 | 0.0152 | 0.5254 | 3 |
| METAB3453 | Physalin L | HMDB0034341 | 0.904 | 0.0155 | 0.5254 | 3 |
| METAB8656 | 5-Hydroxytryptophol glucuronide | HMDB0013200 | -1.368 | 0.0165 | 0.5254 | 3 |
| METAB1653 | NA | NA | -0.714 | 0.0165 | 0.5254 | 3 |
| METAB6203 | D-Tagatose | HMDB0003418 | -0.923 | 0.0167 | 0.5254 | 3 |
| METAB8428 | NA | NA | -1.380 | 0.0169 | 0.5254 | 3 |
| METAB6848 | cis,trans-5'-Hydroxythalidomide | HMDB0013870 | -2.940 | 0.0177 | 0.5254 | 3 |
| METAB5995 | (S)-2-Propylpiperidine | HMDB0030285 | -2.768 | 0.0180 | 0.5254 | 3 |
| METAB3771 | NA | NA | 0.519 | 0.0182 | 0.5254 | 3 |
| METAB2632 | Calcitroic acid | HMDB0006472 | 1.095 | 0.0186 | 0.5254 | 3 |
| METAB5632 | {3-[5-(3,4-dihydroxyphenyl)-6,9,17,19,21-pentahydroxy-18-[3,5,7-trihydroxy-2-(3-hydroxyphenyl)-3,4-dihydro-2H-1-benzopyran-4-yl]-4,12,14-trioxapentacyclohenicosa-2(11),3(8),9,15(20),16,18-hexaen-13-yl]phenyl}oxidanesulfonic acid | HMDB0125940 | -1.046 | 0.0190 | 0.5254 | 3 |
| METAB3426 | DG(14:1(9Z)/15:0/0:0) | HMDB0007039 | -0.900 | 0.0190 | 0.5254 | 3 |
| METAB1822 | Juzirine | HMDB0033954 | -1.360 | 0.0193 | 0.5254 | 3 |
| METAB1021 | NA | NA | -0.727 | 0.0194 | 0.5254 | 3 |
| METAB441 | NA | NA | 1.009 | 0.0195 | 0.5254 | 3 |
| METAB2499 | Prostaglandin F1a | HMDB0002685 | 2.174 | 0.0201 | 0.5254 | 3 |
| METAB1064 | Varenicline | HMDB0015398 | 1.397 | 0.0204 | 0.5254 | 3 |
| METAB9184 | NA | NA | -3.126 | 0.0208 | 0.5254 | 3 |
| METAB9064 | Garcimangosone C | HMDB0036984 | -1.936 | 0.0208 | 0.5254 | 3 |
| METAB9067 | Leonoside B | HMDB0040343 | -1.871 | 0.0209 | 0.5254 | 3 |
| METAB6220 | 2-Hepteneoylglycine | HMDB0094728 | -1.779 | 0.0209 | 0.5254 | 3 |
| METAB8946 | NA | NA | -1.903 | 0.0215 | 0.5254 | 3 |
| METAB2484 | NA | NA | 0.748 | 0.0219 | 0.5254 | 3 |
| METAB3845 | NA | NA | 1.256 | 0.0220 | 0.5254 | 3 |
| METAB6712 | NA | NA | -1.570 | 0.0221 | 0.5254 | 3 |
| METAB415 | Fentanyl | HMDB0014951 | 0.815 | 0.0225 | 0.5254 | 3 |
| METAB6340 | Isoleucylproline | HMDB0011174 | -4.148 | 0.0232 | 0.5254 | 3 |
| METAB483 | Garcimangosone C | HMDB0036984 | 0.866 | 0.0232 | 0.5254 | 3 |
| METAB3274 | NA | NA | 0.586 | 0.0233 | 0.5254 | 3 |
| METAB7602 | Sulforidazine | HMDB0042015 | -4.567 | 0.0234 | 0.5254 | 3 |
| METAB7856 | (3-{5,7-dihydroxy-3-[(3,4,5-trihydroxyoxan-2-yl)oxy]-5H-chromen-2-yl}phenyl)oxidanesulfonic acid | HMDB0127924 | -4.741 | 0.0235 | 0.5254 | 3 |
| METAB8213 | Isomelitric acid A | HMDB0039523 | -2.204 | 0.0235 | 0.5254 | 3 |
| METAB6814 | Praziquantel | HMDB0015191 | -3.715 | 0.0237 | 0.5254 | 3 |
| METAB5999 | Polypropylene glycol (m w 1,200-3,000) | HMDB0032478 | -4.008 | 0.0238 | 0.5254 | 3 |
| METAB9072 | 5,10-Methylenetetrahydromethanopterin | HMDB0060401 | -2.714 | 0.0238 | 0.5254 | 3 |
| METAB6127 | Ethyl 1-(propylthio)propyl disulfide | HMDB0033048 | -1.345 | 0.0239 | 0.5254 | 3 |
| METAB288 | Threoninyl-Arginine | HMDB0029055 | 1.116 | 0.0240 | 0.5254 | 3 |
| METAB213 | Fluvastatin | HMDB0015227 | 3.080 | 0.0240 | 0.5254 | 3 |
| METAB7714 | NA | NA | -1.249 | 0.0248 | 0.5254 | 3 |
| METAB1377 | (S)-N-(4,5-Dihydro-1-methyl-4-oxo-1H-imidazol-2-yl)alanine | HMDB0034912 | 1.230 | 0.0249 | 0.5254 | 3 |
| METAB6117 | 4-Acetamido-2-amino-6-nitrotoluene | HMDB0060384 | -1.613 | 0.0252 | 0.5254 | 3 |
| METAB5738 | xi-4,5-Dihydro-2,4(5)-dimethyl-1H-imidazole | HMDB0037787 | -1.348 | 0.0259 | 0.5254 | 3 |
| METAB9061 | 7,14-dihydroxy-6-methoxy-4,12-dimethyl-5-(3-methylbutanoyl)-10-oxo-2,9-dioxatricyclopentadeca-1(11),3(8),4,6,12,14-hexaene-15-carbaldehyde | HMDB0139003 | -2.073 | 0.0262 | 0.5254 | 3 |
| METAB4823 | NA | NA | -0.699 | 0.0265 | 0.5254 | 3 |
| METAB7211 | NA | NA | -2.051 | 0.0267 | 0.5254 | 3 |
| METAB6482 | 3-Hydroxy-6,8-dimethoxy-7(11)-eremophilen-12,8-olide | HMDB0040756 | -2.484 | 0.0268 | 0.5254 | 3 |
| METAB2079 | Thonzylamine | HMDB0240222 | -0.749 | 0.0269 | 0.5254 | 3 |
| METAB2338 | 6,9,12,15,18,21-Tetracosahexaenoic acid | HMDB0013025 | -0.976 | 0.0272 | 0.5254 | 3 |
| METAB5633 | Tragopogonsaponin K | HMDB0037925 | -1.009 | 0.0276 | 0.5254 | 3 |
| METAB8819 | PE-NMe(14:0/18:3(6Z,9Z,12Z)) | HMDB0112980 | -3.017 | 0.0278 | 0.5254 | 3 |
| METAB440 | NA | NA | 1.019 | 0.0280 | 0.5254 | 3 |
| METAB8270 | NA | NA | -1.271 | 0.0283 | 0.5254 | 3 |
| METAB261 | NA | NA | 1.313 | 0.0283 | 0.5254 | 3 |
| METAB9075 | Aquifoliunine EI | HMDB0032858 | -3.683 | 0.0288 | 0.5254 | 3 |
| METAB3135 | 3-Hydroxy-9-hexadecenoylcarnitine | HMDB0013333 | 0.562 | 0.0290 | 0.5254 | 3 |
| METAB8651 | 6-(1-hydroxy-2-methylbut-3-en-2-yl)-2-(2-hydroxypropan-2-yl)-2H,3H,7H-furo[3,2-g]chromen-7-one | HMDB0130293 | -1.875 | 0.0291 | 0.5254 | 3 |
| METAB3978 | Withaperuvin D | HMDB0030116 | 1.110 | 0.0293 | 0.5254 | 3 |
| METAB8000 | NA | NA | -5.139 | 0.0297 | 0.5271 | 3 |
| METAB484 | Leonoside B | HMDB0040343 | 0.975 | 0.0301 | 0.5290 | 3 |
| METAB299 | Agavoside A | HMDB0034391 | 1.097 | 0.0308 | 0.5355 | 3 |
| METAB1839 | (2S)-2-hydrazinyl-3-(4-hydroxy-3-methoxyphenyl)-2-methylpropanoic acid | HMDB0142637 | 0.606 | 0.0312 | 0.5378 | 3 |
| METAB5396 | TG(14:0/22:6(4Z,7Z,10Z,13Z,16Z,19Z)/o-18:0) | HMDB0042930 | 1.171 | 0.0322 | 0.5449 | 3 |
| METAB7942 | NA | NA | -1.340 | 0.0328 | 0.5449 | 3 |
| METAB7819 | 3-Cyclohexyldodecane | HMDB0036345 | -0.733 | 0.0334 | 0.5449 | 3 |
| METAB454 | [2-hydroxy-2-(5,11,12,13-tetrahydroxy-8-oxo-10-{6,7,13,14-tetrahydroxy-3,10-dioxo-2,9-dioxatetracyclohexadeca-1(15),4,6,8(16),11,13-hexaen-5-yl}-3,7-dioxatricyclotrideca-1(13),9,11-trien-4-yl)ethoxy]sulfonic acid | HMDB0129511 | 0.873 | 0.0337 | 0.5449 | 3 |
| METAB6243 | Dodecanol | HMDB0011626 | -3.802 | 0.0338 | 0.5449 | 3 |
| METAB7067 | Cholesterol | HMDB0000067 | -1.650 | 0.0341 | 0.5449 | 3 |
| METAB6131 | Homoarecoline | HMDB0038321 | -2.453 | 0.0341 | 0.5449 | 3 |
| METAB8660 | NA | NA | -1.086 | 0.0343 | 0.5449 | 3 |
| METAB2345 | cis-4-Decenoic acid | HMDB0004980 | 1.001 | 0.0349 | 0.5449 | 3 |
| METAB4171 | Hydroxydestruxin B | HMDB0040135 | -0.734 | 0.0366 | 0.5449 | 3 |
| METAB524 | Monoethyl malonic acid | HMDB0000576 | -2.247 | 0.0367 | 0.5449 | 3 |
| METAB981 | cis-4-Decenedioic acid | HMDB0000603 | -0.598 | 0.0372 | 0.5449 | 3 |
| METAB7520 | Methacycline | HMDB0015066 | -5.023 | 0.0372 | 0.5449 | 3 |
| METAB6076 | N-Acetylcadaverine | HMDB0002284 | -4.416 | 0.0372 | 0.5449 | 3 |
| METAB4945 | NA | NA | -0.479 | 0.0375 | 0.5449 | 3 |
| METAB8418 | Sorafenib beta-D-Glucuronide | HMDB0060872 | -4.942 | 0.0377 | 0.5449 | 3 |
| METAB3484 | Maraviroc | HMDB0015584 | 0.949 | 0.0378 | 0.5449 | 3 |
| METAB442 | PA(18:1(11Z)/22:2(13Z,16Z)) | HMDB0114914 | 1.015 | 0.0379 | 0.5449 | 3 |
| METAB5910 | 3-Hydroxydodecanoic acid | HMDB0000387 | -1.977 | 0.0381 | 0.5449 | 3 |
| METAB432 | Sorgoleone 358 | HMDB0033443 | 0.950 | 0.0384 | 0.5449 | 3 |
| METAB3212 | Mitoxantrone | HMDB0015335 | -0.640 | 0.0384 | 0.5449 | 3 |
| METAB7908 | Gentamicin | HMDB0014936 | -3.853 | 0.0386 | 0.5449 | 3 |
| METAB1627 | Falcarindiol | HMDB0033941 | 1.002 | 0.0395 | 0.5449 | 3 |
| METAB2860 | 1-Hexanol arabinosylglucoside | HMDB0031689 | -0.239 | 0.0396 | 0.5449 | 3 |
| METAB6475 | 1,3,5-Trichloro-2-methoxybenzene | HMDB0029643 | -1.570 | 0.0400 | 0.5449 | 3 |
| METAB6955 | Capsiate | HMDB0034780 | -5.480 | 0.0402 | 0.5449 | 3 |
| METAB431 | NA | NA | 0.904 | 0.0405 | 0.5449 | 3 |
| METAB8004 | NA | NA | -0.745 | 0.0406 | 0.5449 | 3 |
| METAB419 | 2-amino-4-({1-[(carboxymethyl)-C-hydroxycarbonimidoyl]-2-({1-hydroxy-3-[2-(2-hydroxypropan-2-yl)-7-oxo-2H,3H,7H-furo[3,2-g]chromen-6-yl]-3-methylbutan-2-yl}sulfanyl)ethyl}-C-hydroxycarbonimidoyl)butanoic acid | HMDB0130317 | 1.031 | 0.0409 | 0.5449 | 3 |
| METAB7267 | 2-[Octahydro-4,7-dimethyl-1-oxocyclopenta[c]pyran-3-yl]nepetalactam | HMDB0038596 | -3.000 | 0.0412 | 0.5449 | 3 |
| METAB8985 | [3-(3,5,6,7-tetrahydroxy-4-oxo-4H-chromen-2-yl)phenyl]oxidanesulfonic acid | HMDB0125318 | -1.718 | 0.0412 | 0.5449 | 3 |
| METAB2870 | NA | NA | -0.610 | 0.0418 | 0.5487 | 3 |
| METAB1016 | (Â±)-2-Butylthiazolidine | HMDB0040116 | -0.882 | 0.0424 | 0.5487 | 3 |
| METAB1812 | 2-(3-Carboxy-3-aminopropyl)-L-histidine | HMDB0011655 | -0.586 | 0.0431 | 0.5487 | 3 |
| METAB8477 | Sorafenib beta-D-Glucuronide | HMDB0060872 | -2.931 | 0.0432 | 0.5487 | 3 |
| METAB3825 | PA(10:0/13:0) | HMDB0114773 | 1.242 | 0.0433 | 0.5487 | 3 |
| METAB8322 | Clofarabine | HMDB0014769 | -4.046 | 0.0434 | 0.5487 | 3 |
| METAB528 | Arecoline | HMDB0030353 | -2.115 | 0.0442 | 0.5510 | 3 |
| METAB3518 | NA | NA | -0.714 | 0.0442 | 0.5510 | 3 |
| METAB3863 | N-[(3a,5b,7b)-7-hydroxy-24-oxo-3-(sulfooxy)cholan-24-yl]-Glycine | HMDB0002409 | 2.048 | 0.0445 | 0.5510 | 3 |
| METAB7481 | NA | NA | -3.138 | 0.0458 | 0.5574 | 3 |
| METAB4117 | NA | NA | -0.422 | 0.0461 | 0.5574 | 3 |
| METAB3531 | NA | NA | -0.657 | 0.0472 | 0.5574 | 3 |
| METAB5222 | erythro-6,8-Pentacosanediol | HMDB0041075 | -0.469 | 0.0473 | 0.5574 | 3 |
| METAB250 | NA | NA | 0.919 | 0.0476 | 0.5574 | 3 |
| METAB8478 | NA | NA | -1.391 | 0.0479 | 0.5574 | 3 |
| METAB5987 | NA | NA | -1.890 | 0.0480 | 0.5574 | 3 |
| METAB3576 | 2-({2-[Dimethyl(2-phosphanylethyl)ammonio]ethyl}dimethylammonio)ethyl phosphinite | HMDB0059959 | 1.008 | 0.0483 | 0.5574 | 3 |
| METAB1705 | N4-Acetylcytidine | HMDB0005923 | -0.629 | 0.0483 | 0.5574 | 3 |
| METAB1713 | Acetyl-N-formyl-5-methoxykynurenamine | HMDB0004259 | -1.344 | 0.0485 | 0.5574 | 3 |
| METAB6937 | Carotamine | HMDB0039955 | -2.098 | 0.0489 | 0.5574 | 3 |
| METAB919 | Hydroxyurea | HMDB0015140 | 0.407 | 0.0490 | 0.5574 | 3 |
| METAB7333 | Triton X 100 | HMDB0038380 | -1.501 | 0.0495 | 0.5574 | 3 |
| METAB2326 | 14,15-DiHETrE | HMDB0002265 | 0.864 | 0.0498 | 0.5574 | 3 |
| METAB7402 | NA | NA | -1.821 | 0.0498 | 0.5574 | 3 |
